# Factors influencing post-diarrhea growth patterns among children under two years old in rural Bangladesh

**DOI:** 10.1101/2025.05.30.25328635

**Authors:** Jade Lewis, Yiyi Zhou, Jamie Perin, Sampa Dash, Mohammad Ali, Malathi Ram, Farina Naz, Eva Sultana, Bharati Roy, ABM Ali Hasan, Farzana Afroze, David A Sack, Fahmida Tofail, Tahmeed Ahmed, A.S.G. Faruque, Subhra Chakraborty

## Abstract

Undernutrition in children is a major global health issue. Diarrhea exacerbates undernutrition, creating a vicious cycle of infection and malnutrition. This longitudinal study examined growth patterns and influencing factors among 138 children following diarrhea. Children under two seeking care for watery diarrhea at a rural hospital in Bangladesh were enrolled and followed for three months of home-based visits. Anthropometric measurements—length-for-age z-score (LAZ), weight- for-age z-score (WAZ), weight-for-length z-score (WLZ), and mid-upper arm circumference (MUAC) were taken at enrollment and follow-ups. Linear mixed-effects models assessed associations of clinical, sociodemographic, sanitation, hygiene, and food security factors with growth outcomes. Overall, LAZ declined over three months (-0.260, SD: 0.408), while WAZ, WLZ, and MUAC improved (0.100, SD: 0.484; 0.249, SD: 0.725; 0.672 cm, SD: 0.476). Children with malnutrition at enrollment strongly predicted persistent malnutrition (LAZ < -2, WAZ < -2, WLZ < - 2, MUAC < 12.5 cm) during follow-up compared to new incidence of malnutrition (RR: 3.8, p < 0.001, 95% CI: 1.80, 6.35). Poor growth outcomes were linked to longer diarrhea duration, more vomiting, and hospitalization. Hospitalization was associated with declines in LAZ (-0.095, p = 0.009) and MUAC (-0.098, p = 0.030). Children aged 7–12 months had greater declines in WAZ (- 0.109, p = 0.096) and WLZ (-0.181, p = 0.113). Improved sanitation and hygiene supported growth, while household hunger was linked to declines. This study highlights the multifactorial nature of post-diarrhea growth outcomes and emphasizes the need for integrated interventions addressing clinical care, sanitation, hygiene, and food security.

**Importance:** Children under two years of age in low and middle-income countries are highly vulnerable to undernutrition, especially following diarrheal illness. This study shows that growth recovery after diarrhea is not guaranteed and is shaped by multiple factors, including the child’s age, the severity of illness, household sanitation and hygiene practices, and food security. Children who were already malnourished at the time of illness were much more likely to remain so months later. Poor hygiene practices and limited access to protein-rich foods further reduced the likelihood of recovery. These findings are critical because they highlight the need for targeted follow-up and integrated interventions to support children after diarrhea. By identifying which children are at highest risk and which environmental and clinical conditions hinder growth, this study provides essential evidence to guide strategies for improving child nutrition and health in similar low-resource settings.

## 1 Introduction

Despite remarkable progress towards the 2025 Global Nutrition Targets[1] and 2030 Sustainable Development Goals (SDG) 2.2[2] in the past few decades, child undernutrition remains a significant global health challenge, disproportionately affecting children under 5 years old in low- and middle-income countries (LMICs)[3]. According to recent global estimates in 2020 and 2022, 148.1 million (22.3%) children under five years old suffered from stunting, 45.0 million (6.8%) from wasting, and 85.4 million (12.6%) from underweight[4,5]. Notably, 95% of the stunted children and 97% of the wasted children were in Asia and Africa[4]. Accumulated evidence indicates that undernutrition in early life is associated with a higher risk of premature deaths related to infections such as diarrhea, measles, and pneumonia, accounting for 45% of child mortality, and impairs neurobehavioral and cognitive development among those survivors[6–8].

Bangladesh, as a South Asian country, bears one of the highest burdens of child undernutrition. The country has initiated the first and second National Plans of Action for Nutrition, which outlined strategies to eliminate malnutrition in its population. It has achieved impressive declines in childhood undernutrition, as can be seen from a decrease of 50.3% in the prevalence of stunting and another decrease of 38.3% in the prevalence of underweight among children under five between 1991 and 2022[9–11]. Yet, with 23.6%, 22.3%, and 11% of the children continue to suffer from stunting, underweight, and wasting, respectively, according to the most recent national survey in 2022[9–11].Undernutrition continues to contribute significantly to child mortality, having been identified as an underlying factor in up to 60% of deaths among children under five, primarily by increasing vulnerability to infectious diseases and impaired immune function..[13].

Determinants of child undernutrition could be multifold, including food insecurity and poor nutrition, maternal factors (e.g., low maternal education level, poor maternal nutrition, short maternal stature, etc.), household factors (e.g., low household income, size of the family, unimproved sanitation and water supply), inadequate breastfeeding and complementary feeding practices, low birth weight, and severe infections early in life, which varies by population, sites, and demographic factors[14–19].

Although the etiology of child undernutrition is complex, symptomatic and asymptomatic infections by common enteric pathogens have consistently been a pivotal contributor to child undernutrition in LMICs[6,20,21]. A comparative risk assessment analysis across 137 developing countries estimated that diarrhea was responsible for 5.8 million (13.1%) stunted children worldwide[16,20].

While WASH interventions consistently reduce diarrheal incidence in LMICs [57, 58], their impact on linear growth remains equivocal. Large, randomized trials – including the WASH Benefits Bangladesh trial [59] and the SHINE trial in Zimbabwe [60] – found that even combined water, sanitation, handwashing, and nutrition interventions had only marginal effects on stunting, despite significant reductions in diarrhea. A mediation analysis of the WASH Benefits Bangladesh trial further revealed that improved sanitation reduced biomarkers of environmental enteric dysfunction but failed to substantially improve linear growth [56], suggesting that non-diarrhea pathways (e.g., chronic inflammation, nutrient malabsorption) may dominate the causal pathway. The MAL-ED study corroborated this, demonstrating that diarrheal burden alone explained less than 5% of growth faltering across diverse settings [61]. These findings underscore the need to examine post-diarrheal recovery in context-specific ways, including factors like microbiota composition or repeated subclinical infections [62]. The close link between diarrhea and undernutrition is concerning as undernourished children are more susceptible to gastrointestinal infections and diarrhea, thus falling into a diarrhea-undernutrition-diarrhea cycle [23]. In rural Bangladesh, where this cycle persists despite WASH improvements [59,61], identifying factors that disrupt it – particularly those influencing post-diarrhea growth trajectories – could unlock targeted strategies to mitigate chronic undernutrition.

It is pivotal to break out of this vicious cycle and improve the chronic sequelae in children who frequently experience diarrhea during their first few years of life. However, evidence is limited in rural Bangladesh on the key factors affecting post-diarrhea undernutrition among children. A previous study found that moderate-to-severe diarrhea, children aged 24-59 months, lower mother’s education, and lower household income were significantly associated with undernutrition in children with diarrhea presenting at the hospital[24]. Another study focused on examining environmental risk factors of wasting in children admitted to a hospital in Bangladesh but did not account for the role of diarrhea[25]. Current evidence related to risk factors of child undernutrition in Bangladesh was mostly generated from cross-sectional analyses, while individual factors may change over time, and so do their relationships with undernutrition[25]. Knowledge of how longitudinal history of diarrhea and other important factors affect children’s growth is still lacking.

In this study, we aim to characterize growth patterns following diarrhea and identify key factors influencing post-diarrhea growth trajectory among children under two years old with all-cause watery diarrhea seeking care from a tertiary level hospital in rural Bangladesh.

## 2 Methods

### 2.1 Study Description and Study Population

#### 2.1.1 Study Site

Mirzapur is a rural sub-district (Upazila) of Bangladesh that covers 374 Square km in Tangail district. It is located 60 km northwest of the capital city, Dhaka. Kumudini Women’s Medical College and Hospital is a 750-bed nonprofit hospital located in the central urban union of Mirzapur was established in 1938 to provide health services to the surrounding poor rural population. Since 1982, a diarrhea treatment unit was created and serves nearly 1,500 diarrhea patients each year for treatment. The Kumudini Hospital provides free treatment to all those who are unable to afford medical care.

#### 2.1.2 Study Description

Children under two years old with any watery diarrhea seeking care at the Kumudini Hospital were enrolled in the ongoing study Impact of Non-Dysentery Shigella Infection on the Growth and Health of Children over Time (INSIGHT) in Bangladesh and followed for three months from enrollment[26].

#### 2.1.3 Inclusion and Exclusion Criteria

The inclusion criteria of the study population were children > 1 month and < 24 months of age seeking care at Kumudini Hospital for watery diarrhea, residing within the catchment area, and willing to be available for sample and data collection during the scheduled visits. Exclusion criteria included diarrhea started more than 48 hours before enrollment, having already taken antibiotics for diarrhea in the past three days or having antibiotics prescribed for current diarrhea, presenting with another significant disease process requiring specific therapy, presenting with severe acute malnutrition (i.e., WLZ < -3) or bilateral oedema. Children who only completed the baseline anthropometry measurements or were withdrawn from the study before the second anthropometry measurement were excluded from the analysis.

### 2.2 Data Collection

Socio-demographic information, diarrhea morbidity data, sanitation and hygiene, and food security information were collected by administering questionnaires to caregivers of the study children at the baseline visit. Diarrhea morbidity data were collected every other day following the baseline visit till one week and then every two days until day 90 via a questionnaire administered to the caregivers. Anthropometry of the children was measured at the baseline, followed by days 14, 30, 60, and 90. Medical records were reviewed to identify any hospitalization for diarrhea illness.

### 2.3 Outcome Assessment

The continuous outcomes of interest were changes in length-for-age z score (LAZ), weight-for-length z score (WLZ), weight-for-age z scores (WAZ), and mid-upper arm circumference (MUAC) between baseline and each follow-up anthropometry measurement. Enrollment anthropometry measurement of WAZ, WLZ, and MUAC occurred after rehydration therapy was given to the children. The binary outcomes of interest were four indicators of undernutrition (MUAC <12.5 cm), including stunting (i.e., LAZ < -2), wasting (WLZ < -2), and underweight (WAZ < -2), using the WHO reference population[7,27]. Anthropometry z scores were calculated based on the WHO Child Growth Standards using the WHO-issued “Anthro” package in R software[28].

### 2.4 Risk Factors

Several clinical, socio-demographic, sanitation and hygiene, and food security factors were examined in this analysis, which was based on WHO and UNICEF conceptual frameworks on child nutrition[29–31] and previous literature[32–34]. The clinical factors included are the severity of diarrhea (based on the Modified Vesikari Severity Score)[57], number of stools in the last 24 hours of enrollment, duration of diarrhea, vomiting and fever since last visit, dehydration status at enrollment (based on the WHO guidelines for dehydration), hospitalization due to diarrhea, and anorexia at enrollment (defined as the loss of appetite).

The sanitation and hygiene factors included are the type of soap found in the household for handwashing (based on whether the field worker visually confirmed soap in the household), reported handwashing before preparing food and feeding children, and toilet facility type. Under toilet facility, improved sanitation was defined as facilities separating human waste from human contact, flush/pour-flush to piped sewer system, ventilated-improved pit latrines, or pit latrines with slab or composting toilets; unimproved sanitation contains shared toilet facilities, flush/pour-flush not to piped sewer system, pit latrines without slabs, bucket or hanging latrines, and open defecation[32].

The food security factors included gathering information using several questions about whether the household couldn’t afford enough food or eat balanced meals that include fish or meat etc. These food security factors were then assigned points to determine the household hunger scale. The scale used in this study was derived from the Household Hunger Scale from the Food and Nutrition Technical Assistance III Project (FANTA)[53]. The score ranged from 0-8 points, where household with 0-1 points being categorized as “little to no hunger in household”, 2-4 points being categorized as “moderate hunger in household”, and 5-8 points being categorized as “severe hunger in household”.

### 2.5 Statistical Analysis

Descriptive statistics of children’s baseline characteristics were computed. The mean and variance of change in the LAZ, WAZ, and WLZ, respectively, relative to the baseline value as well as the prevalence of stunting, wasting, and underweight at each visit were computed and visually displayed for the full cohort and by each subgroup of risk factors in the exploratory analysis. Diarrhea was defined as three or more loose or liquid stools per day, and a new diarrhea episode was defined as onset of diarrhea after three consecutive diarrhea-free days. Spaghetti plots were used to visualize individuals’ trajectories of change in anthropometry z scores and MUAC, and lasagna plots were used to reflect each child’s change in their status of stunting, wasting, underweight, and malnutrition over time. To compare the risk of malnutrition at follow-up between children who were malnourished and those who were not malnourished at enrollment, we constructed a 2×2 contingency table based on malnutrition status at enrollment and at day 90. We used Fisher’s exact test to assess differences in proportions and calculated risk ratios with corresponding 95% confidence intervals.

Bivariate and multivariate linear mixed-effect models were conducted to estimate the average change in anthropometry z scores over follow-up and 95% CIs associated with the risk factors of interest. In the bivariate models, the mean model consisted of a linear term of time and potential risk factors (one at a time), with or without an interaction term between them. Variables with a p-value <0.15 in the bivariate analysis-either in the main effect or its interaction with time-were considered for inclusion in the multivariable model. In building the multivariable model, we considered potential confounders based on conceptual frameworks by WHO/UNICEF and prior literature. These included child age, sex, baseline anthropometric status, maternal education, household income, and household sanitation and hygiene characteristics. Variables were retained in the final model if they remained statistically significant (p<0.05). This approach allowed us to adjust for important structural and behavioral factors potentially influencing child growth. Variance Inflation Factor (VIF) was used to check potential multicollinearity. Regarding the random-effects model, a random intercept-only model, a random-slope-only model, and a random intercept and random slope model were explored sequentially for each potential risk factor, using Maximum Likelihood (ML) for model comparison and using Restricted Maximum Likelihood (REML) for coefficient estimates report. Likelihood ratio test (LRT) was used to determine whether an interaction term of fixed effects was included. The selection of random effects models was based on the Akaike information criterion (AIC) and the Bayesian information criterion (BIC), leading to the random intercept and random slope models appearing to be the best-fitting model for all risk factors. Cluster-robust variance estimation was used to obtain the 95% robust CIs of the coefficient estimates.

Although total diarrhea duration over the follow-up period was analyzed to assess its impact on growth outcomes, no significant associations were found; therefore, these results are not reported, and the following results focus on the index case of diarrhea.

The significance level was set to 0.05 for all tests. Data management and analyses were conducted in R (version 4.3.1, 2023-06-16).

### 2.6 Ethical Considerations

Caregivers of all study participants reviewed and signed informed consent. The institutional review boards of the International Center for Diarrheal Disease Research, Bangladesh (icddr,b) and Johns Hopkins Bloomberg School of Public Health approved the study protocol (IRB number: 00015285).

## 3 Results

### 3.1 Baseline Characteristics of the Participants

A total of 138 children under 24 months of age with watery diarrhea, seeking care at Kumudini Hospital in rural Bangladesh, were included in the analysis. Anthropometric measurements were taken at baseline and included five metrics. The median age at baseline was 9.1 months (IQR: 6.0-14.8), and 60.9% of the children were male (Table 1). More than half of the children (55.8%) came from families earning below 20,000 Taka (167.34 USD) per month. In this population, 68.8% of the household heads had an education level of either illiterate or primary, while 52.9% of mothers had completed secondary or higher education. According to the Household Hunger Scale, 5.1% of the population experienced moderate hunger, while none reported severe hunger.

**Table 1.**
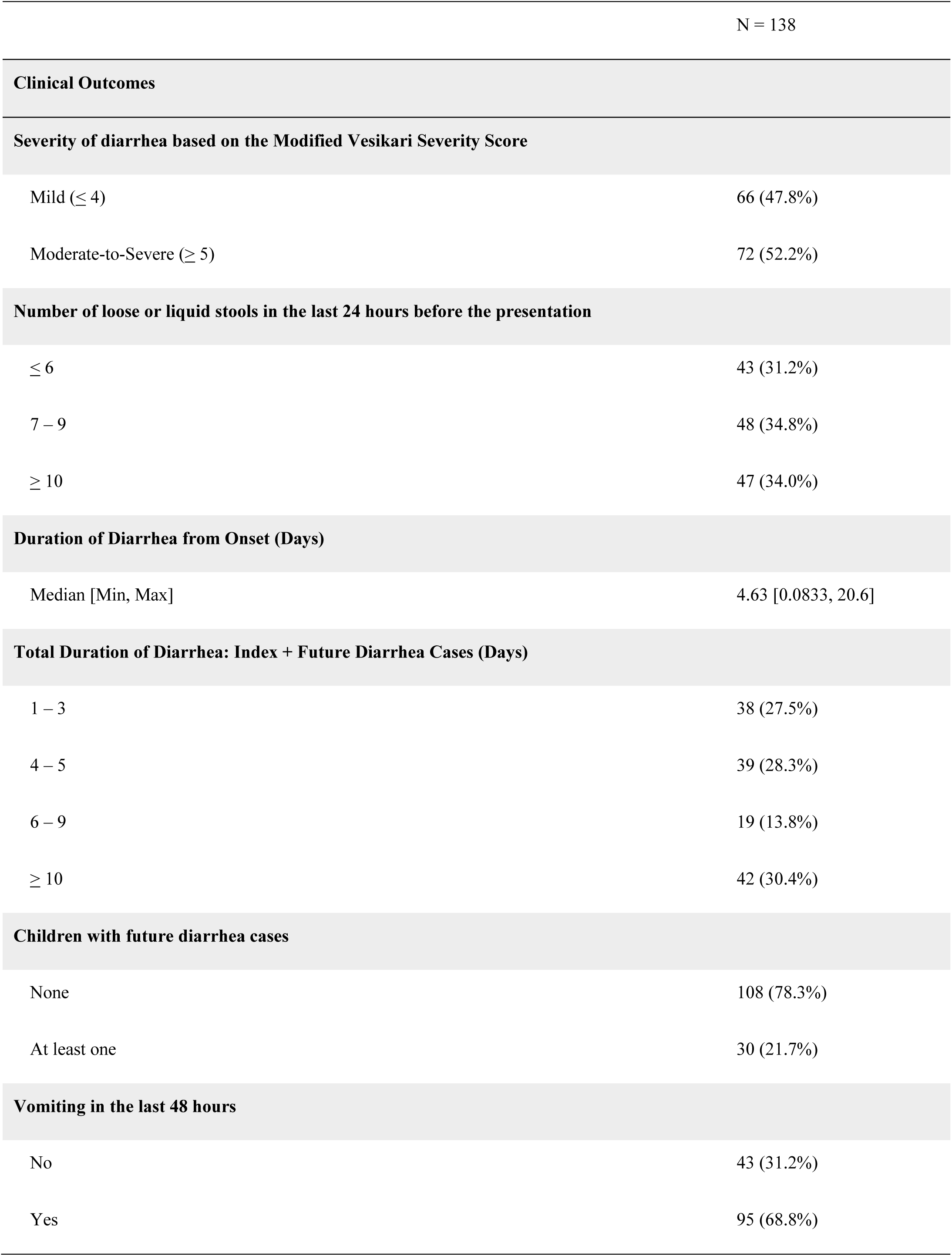
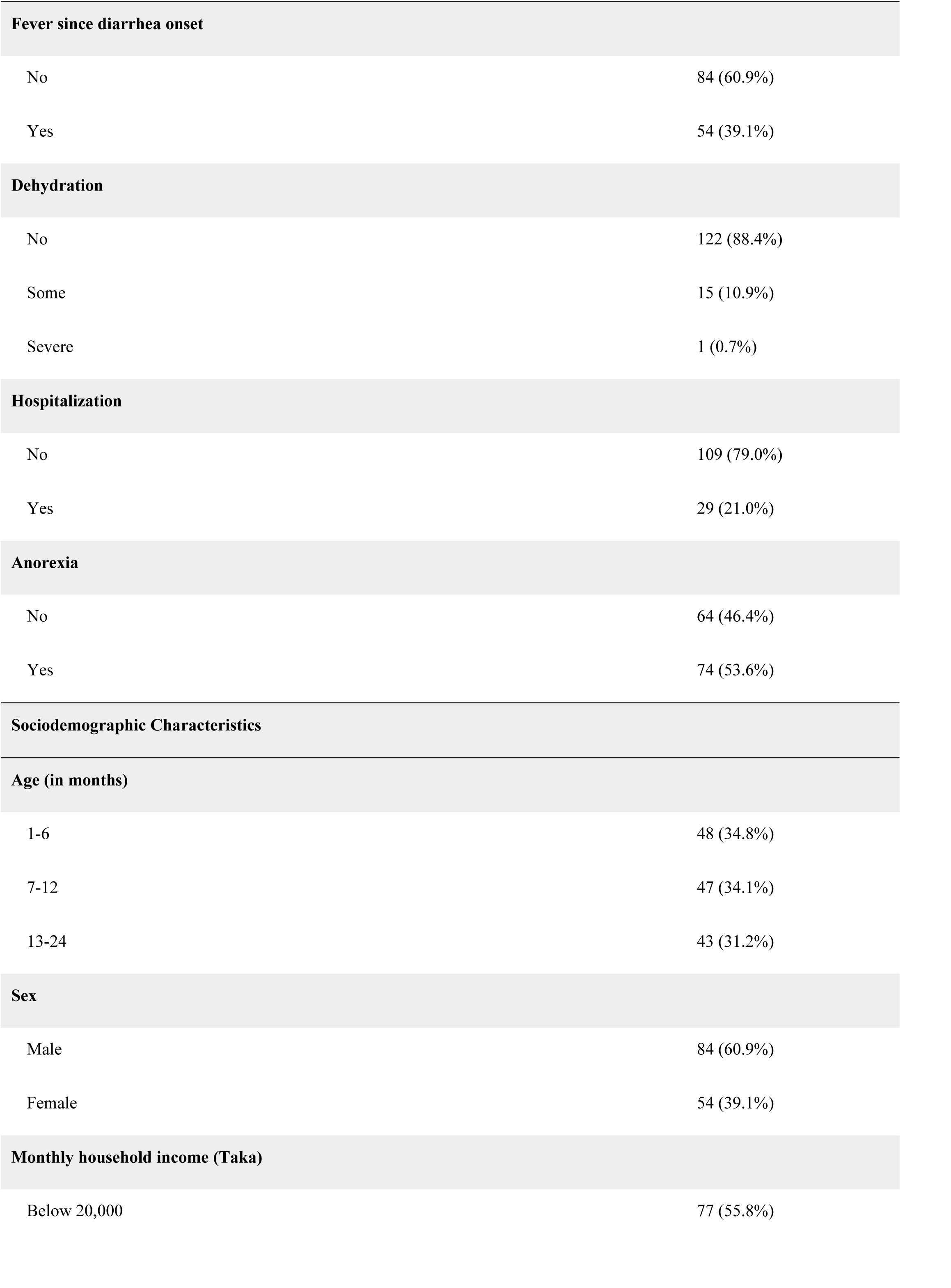
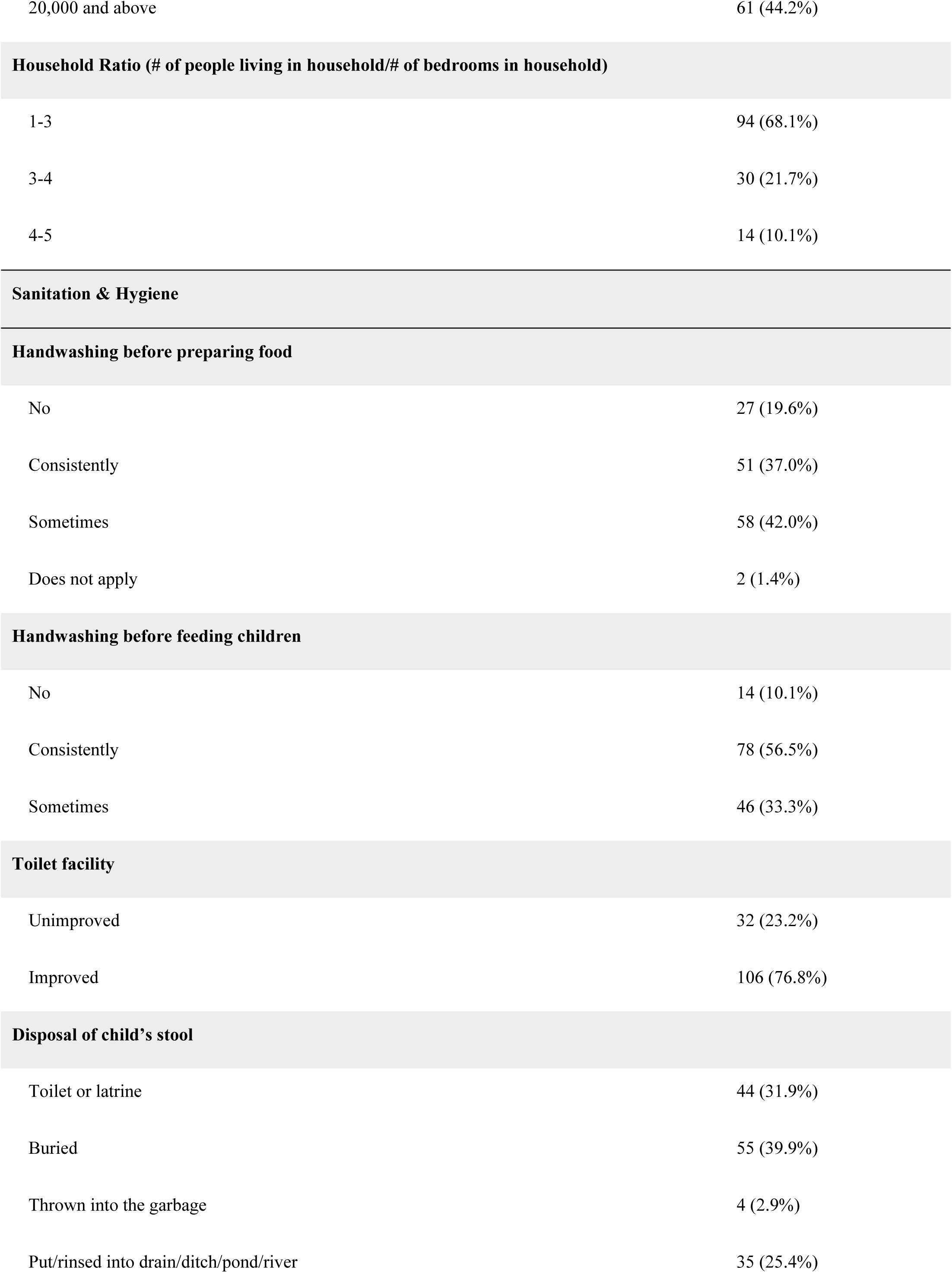
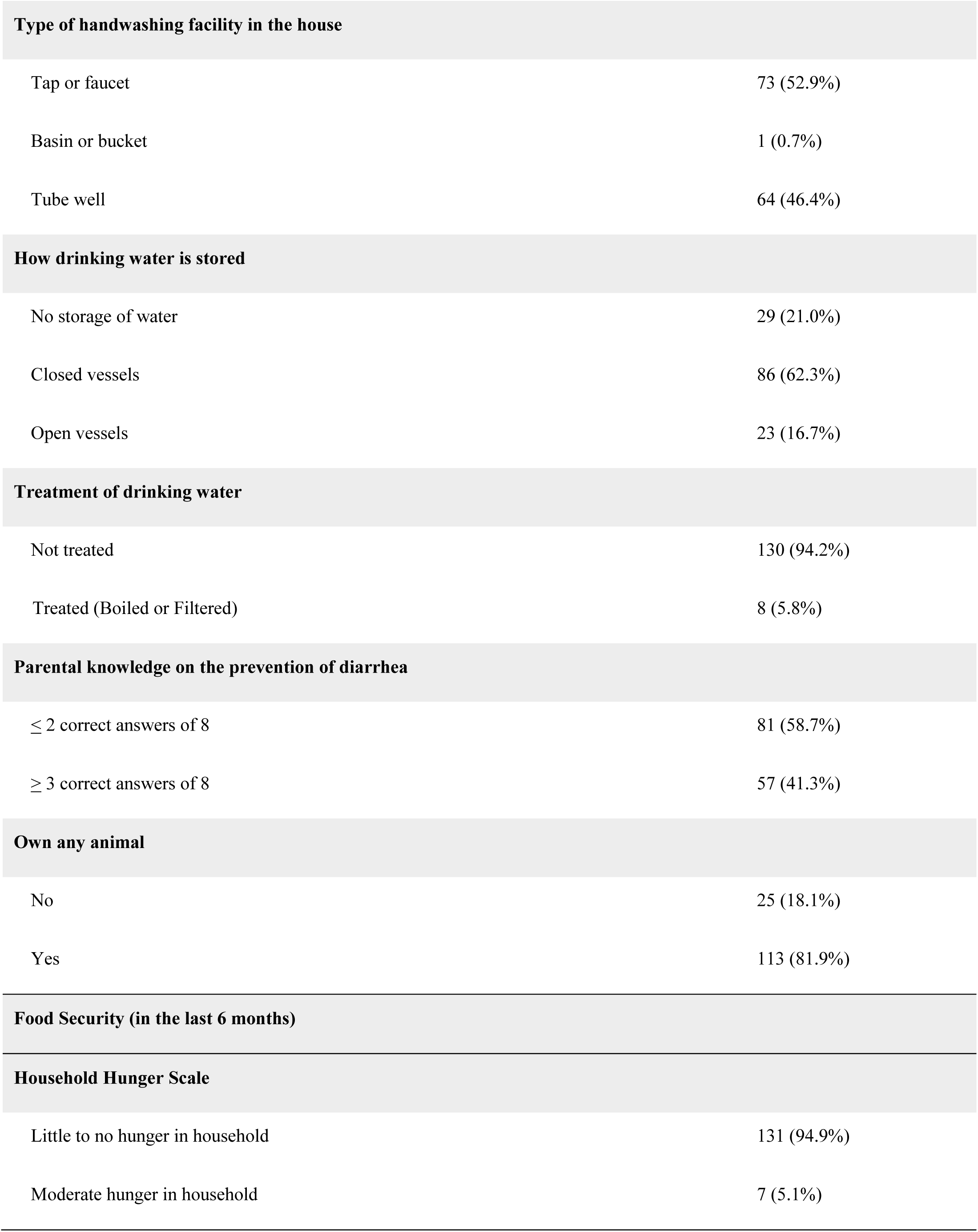
Baseline characteristics of participants.

| N = 138 |  |
| --- | --- |
| **Clinical Outcomes** |  |
| **Severity of diarrhea based on the Modified Vesikari Severity Score** |  |
| Mild ($\leq 4$) | 66 (47.8%) |
| Moderate-to-Severe ($\geq 5$) | 72 (52.2%) |
| **Number of loose or liquid stools in the last 24 hours before the presentation** |  |
| $\leq 6$ | 43 (31.2%) |
| 7 – 9 | 48 (34.8%) |
| $\geq 10$ | 47 (34.0%) |
| **Duration of Diarrhea from Onset (Days)** |  |
| Median [Min, Max] | 4.63 [0.0833, 20.6] |
| **Total Duration of Diarrhea: Index + Future Diarrhea Cases (Days)** |  |
| 1 – 3 | 38 (27.5%) |
| 4 – 5 | 39 (28.3%) |
| 6 – 9 | 19 (13.8%) |
| $\geq 10$ | 42 (30.4%) |
| **Children with future diarrhea cases** |  |
| None | 108 (78.3%) |
| At least one | 30 (21.7%) |
| **Vomiting in the last 48 hours** |  |
| No | 43 (31.2%) |
| Yes | 95 (68.8%) |
| **Fever since diarrhea onset** |  |
| No | 84 (60.9%) |
| Yes | 54 (39.1%) |
| **Dehydration** |  |
| No | 122 (88.4%) |
| Some | 15 (10.9%) |
| Severe | 1 (0.7%) |
| **Hospitalization** |  |
| No | 109 (79.0%) |
| Yes | 29 (21.0%) |
| **Anorexia** |  |
| No | 64 (46.4%) |
| Yes | 74 (53.6%) |
| **Sociodemographic Characteristics** |  |
| **Age (in months)** |  |
| 1-6 | 48 (34.8%) |
| 7-12 | 47 (34.1%) |
| 13-24 | 43 (31.2%) |
| **Sex** |  |
| Male | 84 (60.9%) |
| Female | 54 (39.1%) |
| **Monthly household income (Taka)** |  |
| Below 20,000 | 77 (55.8%) |
| 20,000 and above | 61 (44.2%) |
| Household Ratio (# of people living in household/# of bedrooms in household) |  |
| 1-3 | 94 (68.1%) |
| 3-4 | 30 (21.7%) |
| 4-5 | 14 (10.1%) |
| Handwashing before preparing food |  |
| No | 27 (19.6%) |
| Consistently | 51 (37.0%) |
| Sometimes | 58 (42.0%) |
| Does not apply | 2 (1.4%) |
| Handwashing before feeding children |  |
| No | 14 (10.1%) |
| Consistently | 78 (56.5%) |
| Sometimes | 46 (33.3%) |
| Toilet facility |  |
| Unimproved | 32 (23.2%) |
| Improved | 106 (76.8%) |
| Disposal of child's stool |  |
| Toilet or latrine | 44 (31.9%) |
| Buried | 55 (39.9%) |
| Thrown into the garbage | 4 (2.9%) |
| Put/rinsed into drain/ditch/pond/river | 35 (25.4%) |

| Type of handwashing facility in the house |  |
| --- | --- |
| Tap or faucet | 73 (52.9%) |
| Basin or bucket | 1 (0.7%) |
| Tube well | 64 (46.4%) |
| How drinking water is stored |  |
| No storage of water | 29 (21.0%) |
| Closed vessels | 86 (62.3%) |
| Open vessels | 23 (16.7%) |
| Treatment of drinking water |  |
| Not treated | 130 (94.2%) |
| Treated (Boiled or Filtered) | 8 (5.8%) |
| Parental knowledge on the prevention of diarrhea |  |
| $\leq 2$ correct answers of 8 | 81 (58.7%) |
| $\geq 3$ correct answers of 8 | 57 (41.3%) |
| Own any animal |  |
| No | 25 (18.1%) |
| Yes | 113 (81.9%) |
| Food Security (in the last 6 months) |  |
| Household Hunger Scale |  |
| Little to no hunger in household | 131 (94.9%) |
| Moderate hunger in household | 7 (5.1%) |

Based on the Modified Vesikari Severity score, 52.2% of children had moderate-to-severe diarrhea, and 34.0% experienced more than ten loose stools within 24 hours of hospital presentation. The median duration of diarrhea from enrollment was 4.63 days [0.08, 20.6], and only 10.9% had signs of any dehydration. Vomiting within 48 hours of hospital presentation was reported in 68.8% of cases, fever was present in 39.1%, and 53.6% had anorexia. Approximately 21.0% of children were hospitalized due to their diarrhea episode.

Sanitation and hygiene practices varied among participants. Over half of the households (52.9%) had tap or faucet facilities for handwashing, while 46.4% relied on tube wells. Improved toilet facilities were present in 76.8% of households, while 23.2% had unimproved facilities. Handwashing practices before preparing food were inconsistent, with 37.0% of caregivers reporting consistent handwashing, 42.0% reporting intermittent handwashing, and 19.6% reporting no handwashing. Similarly, 56.5% of caregivers reported consistent handwashing before feeding children, while 33.3% reported intermittent handwashing, and 10.1% reported no handwashing. Drinking water storage practices also varied, with 62.3% of households storing water in closed vessels, 16.7% in open vessels, and 21.0% reporting no storage of water.

### 3.2 Growth Patterns After Diarrhea and Prevalence of Undernutrition Over Time

The population-average change in length-for-age z-score (LAZ) showed a linear decrease over the three-month follow-up period (Figure 1A), while weight-for-age z-score (WAZ), weight-for-length z-score (WLZ), and mid-upper arm circumference (MUAC) exhibited a linear increase (Figures 1B, 1C, & 1D). There was notable individual variation in the growth trajectories. By the end of the 90-day follow-up, the average changes in z-scores were -0.260 (SD: 0.408) for LAZ, 0.100 (SD: 0.484) for WAZ, 0.249 (SD: 0.725) for WLZ, and MUAC increased by an average of 0.672 cm (SD: 0.476).

**Figure 1.**
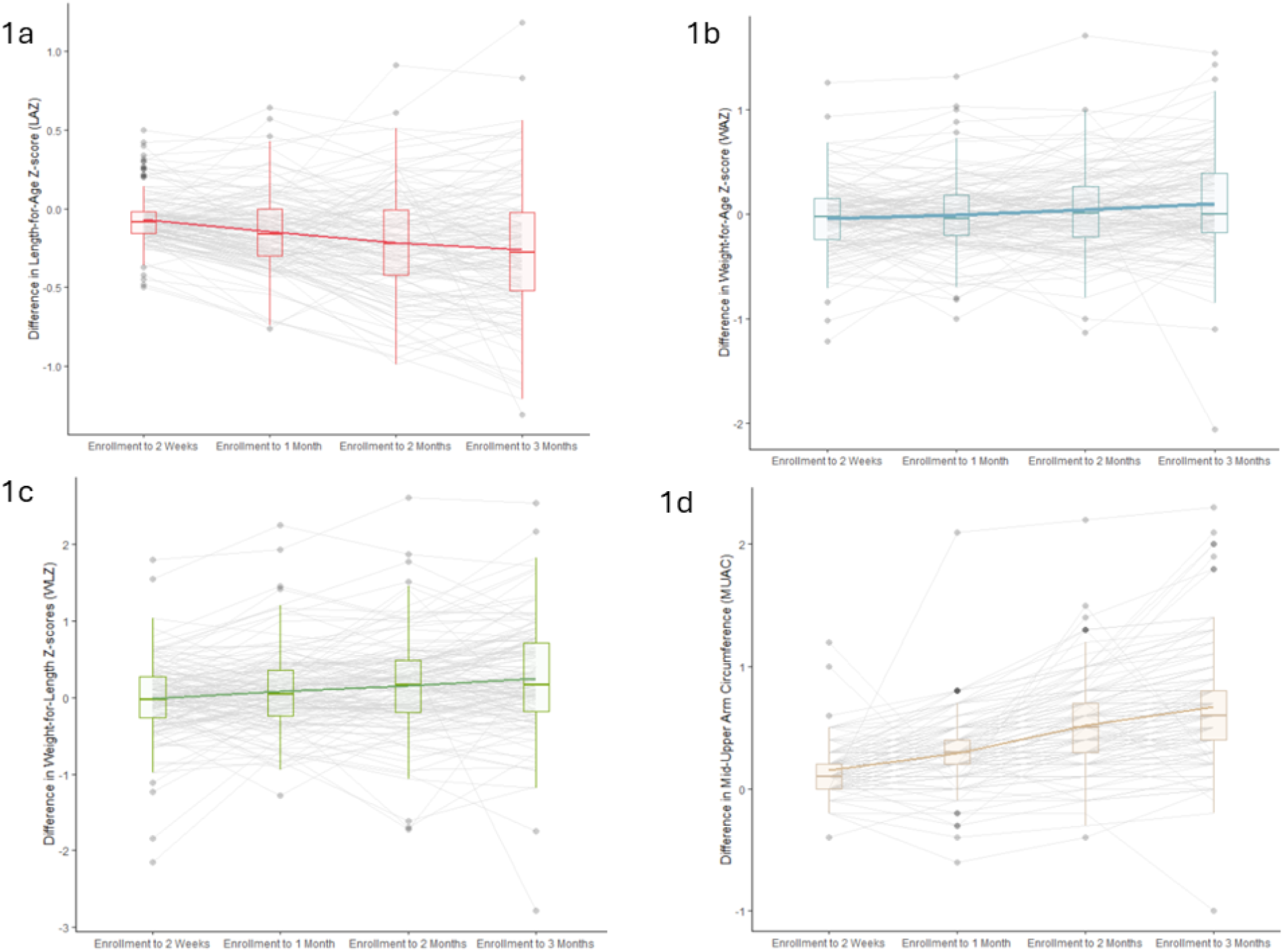
The population average change in Length-for-Age Z-score (a), Weight-for-Age Z-score (b), Weight-for-Length Z-score (c), and Mid-Upper Arm Circumference (MUAC) over the three-month follow-up period with individual growth trajectories.

Although the growth status changed at the individual level for some children, overall, there were no significant changes in the prevalence of stunting, wasting, or undernutrition, as determined by LAZ, WAZ, WLZ, and MUAC scores, over the observation period (Figure 2a-2d). At enrollment, 12 children (8.7%) were stunted, six (4.3%) were wasted, 13 (9.4%) were underweight, and eight (5.8%) had a MUAC ≤12.5 cm, indicating undernutrition. By day 90, four of the stunted children, two of the wasted children, and three of the underweight children had recovered, while the remaining children in each group continued to show deficits. All eight children with low MUAC at enrollment had recovered by day 90. At follow-up, 12 children were stunted—eight who were stunted at baseline and four newly stunted. Similarly, 12 children were underweight at day 90, including 10 who were underweight at enrollment and two new cases. Seven children were wasted at follow-up; four were persistently wasted, and three became newly wasted.

**Figure 2.**
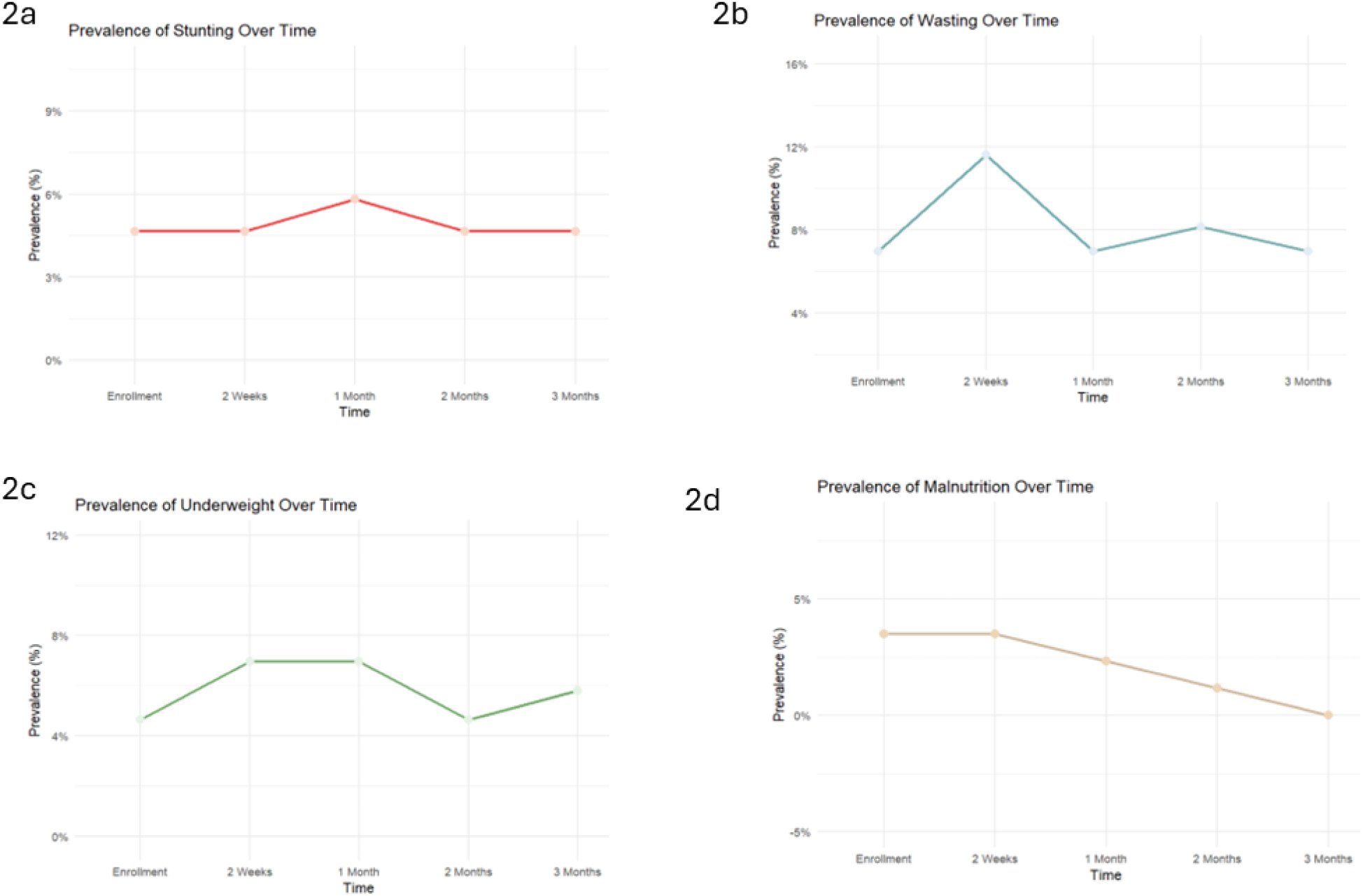
Prevalence of stunting (a), wasting (b), underweight (c), and malnutrition (d) in the study population over the three-month follow-up period post enrollment.

Children who were malnourished at enrollment were significantly more likely to remain malnourished at follow-up than those who were well-nourished at baseline. Of the 25 children who met at least one malnutrition criterion (stunted, wasted, or underweight) at enrollment, 18 (72%) remained malnourished at day 90. In contrast, only 6 of the 113 children (5.3%) who were not malnourished at enrollment developed a new nutritional deficit by follow-up. This corresponds to a risk ratio of 3.8 (p<0.001, 95% CI: 1.80, 6.35), indicating a significantly higher risk of persistent or incident malnutrition among children who were malnourished at baseline.

### 3.3 Factors Associated with Change in Growth Parameters

Tables 2 and Supplementary Tables 1a-1d present the results of bivariate and multivariate linear mixed-effects models assessing the association between changes in WAZ, LAZ, WLZ, and MUAC and sociodemographic, sanitation, hygiene, food security, and clinical risk factors.

**Table 2.**
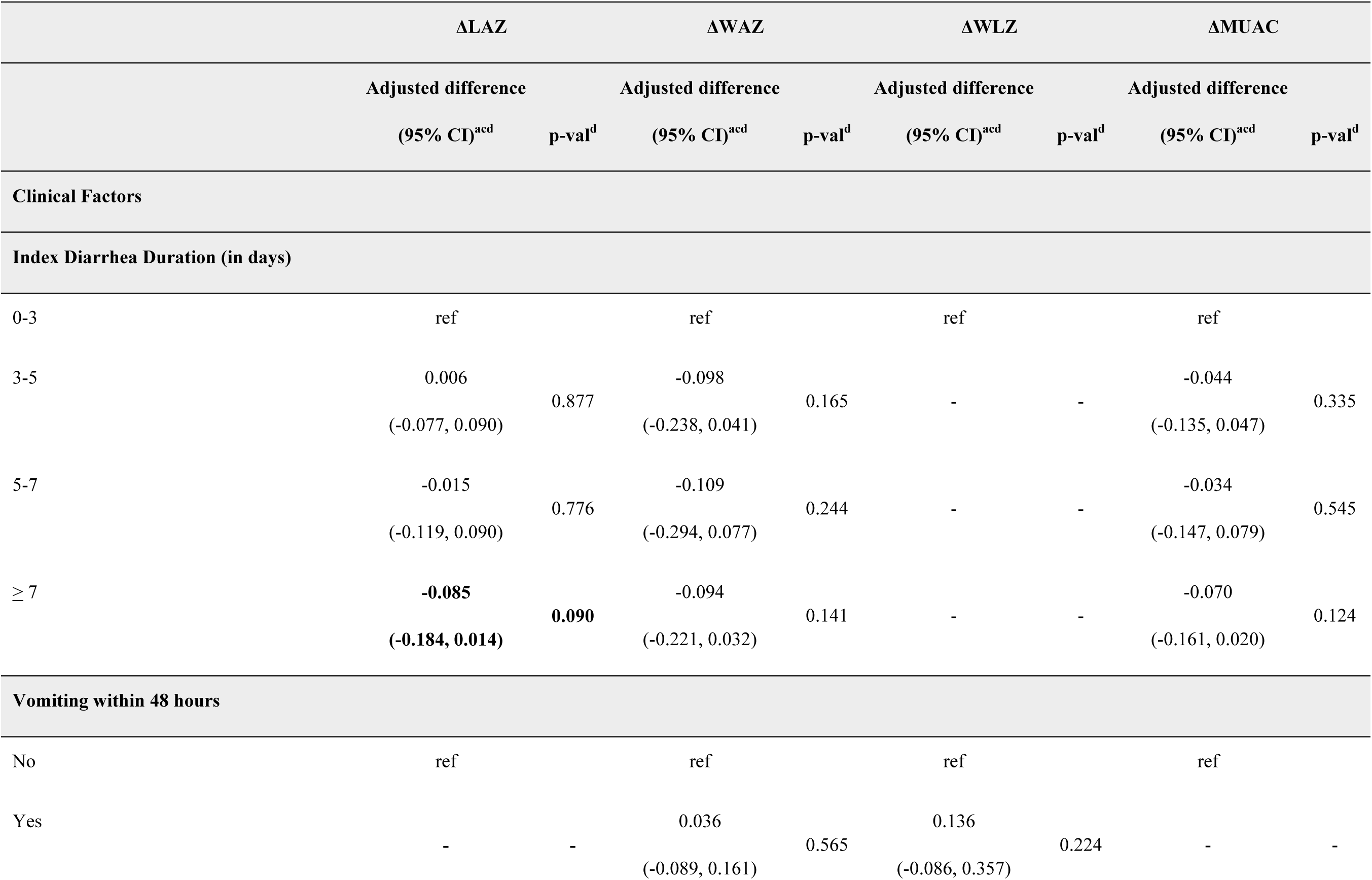
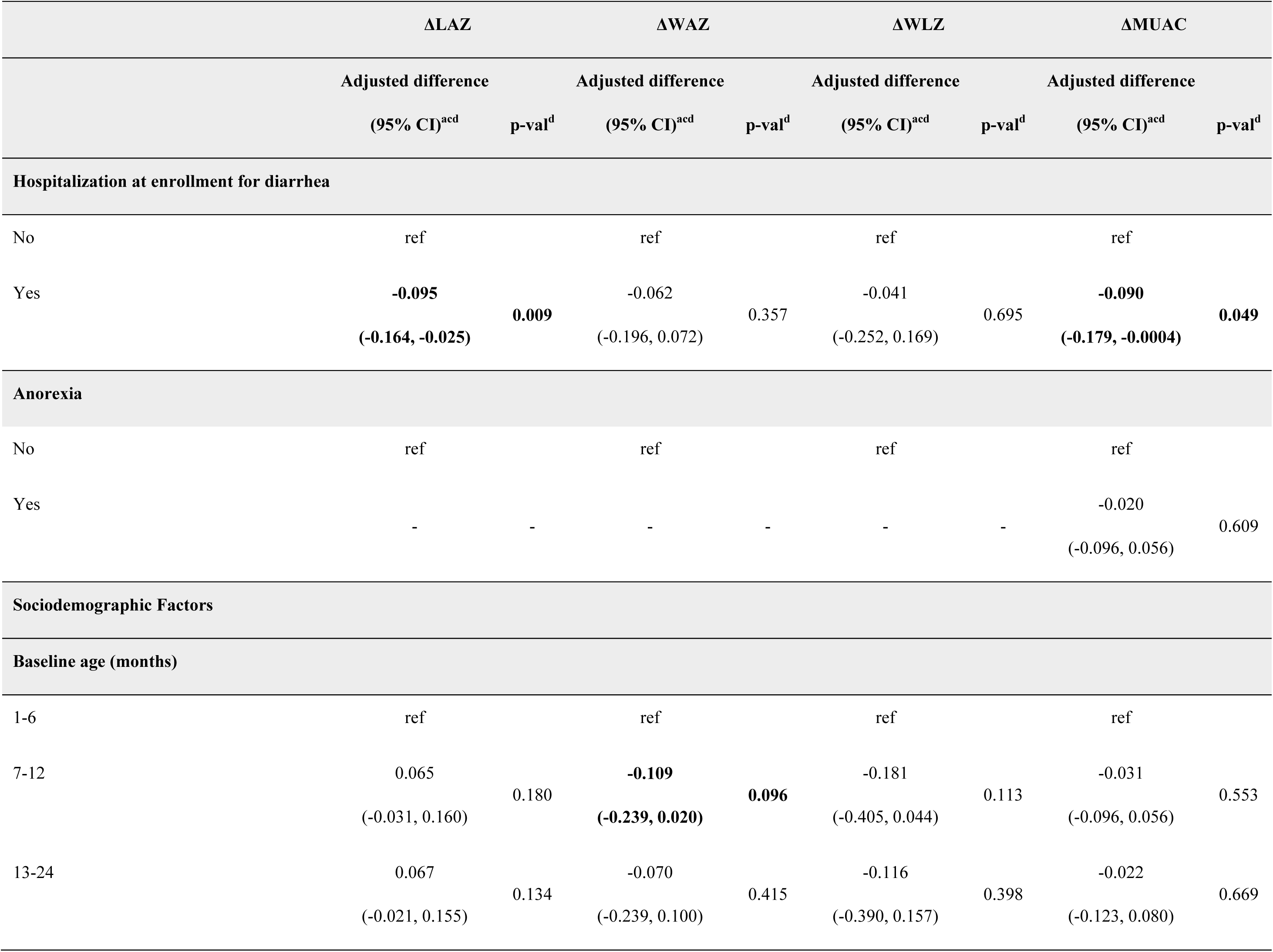
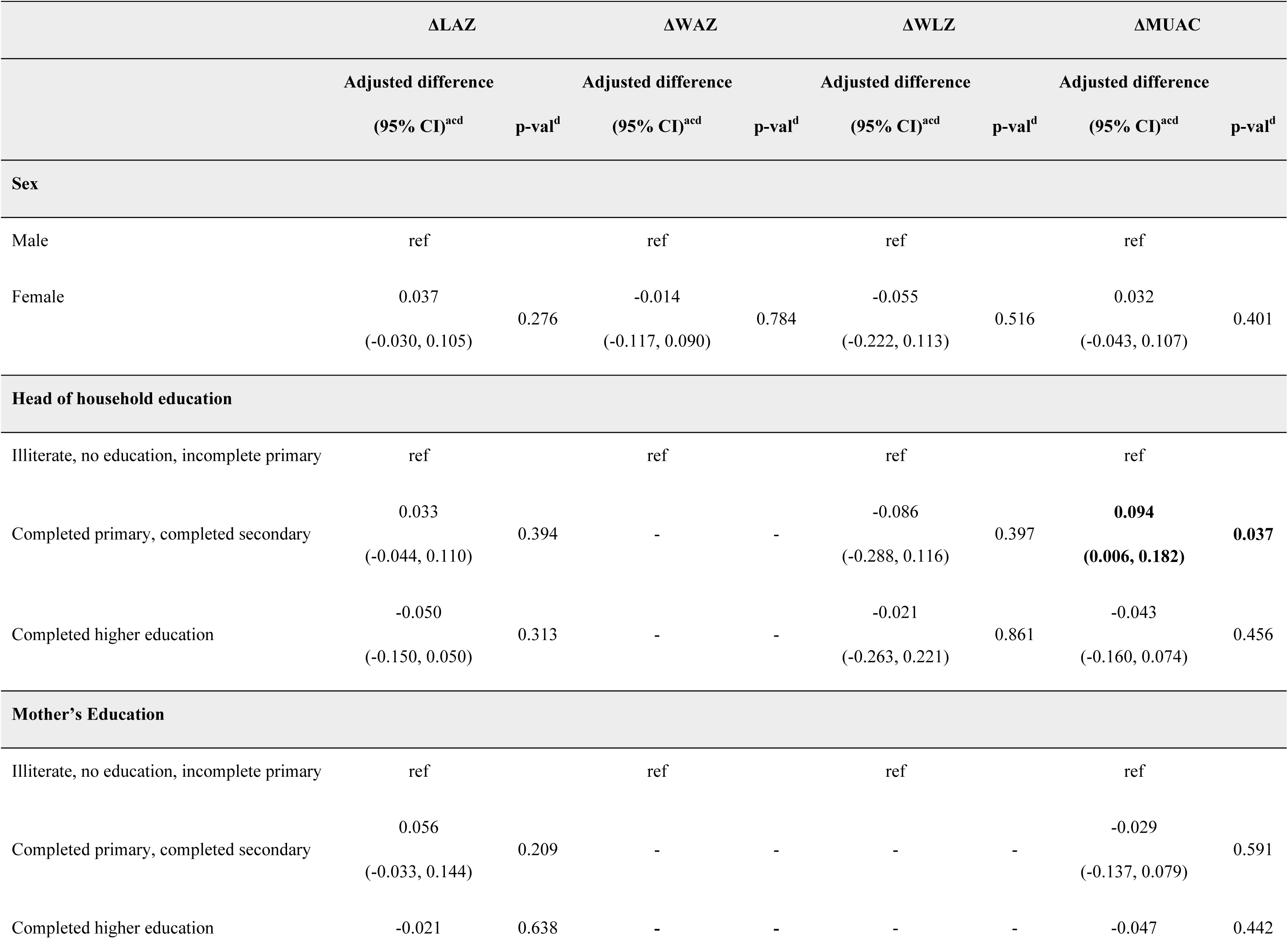
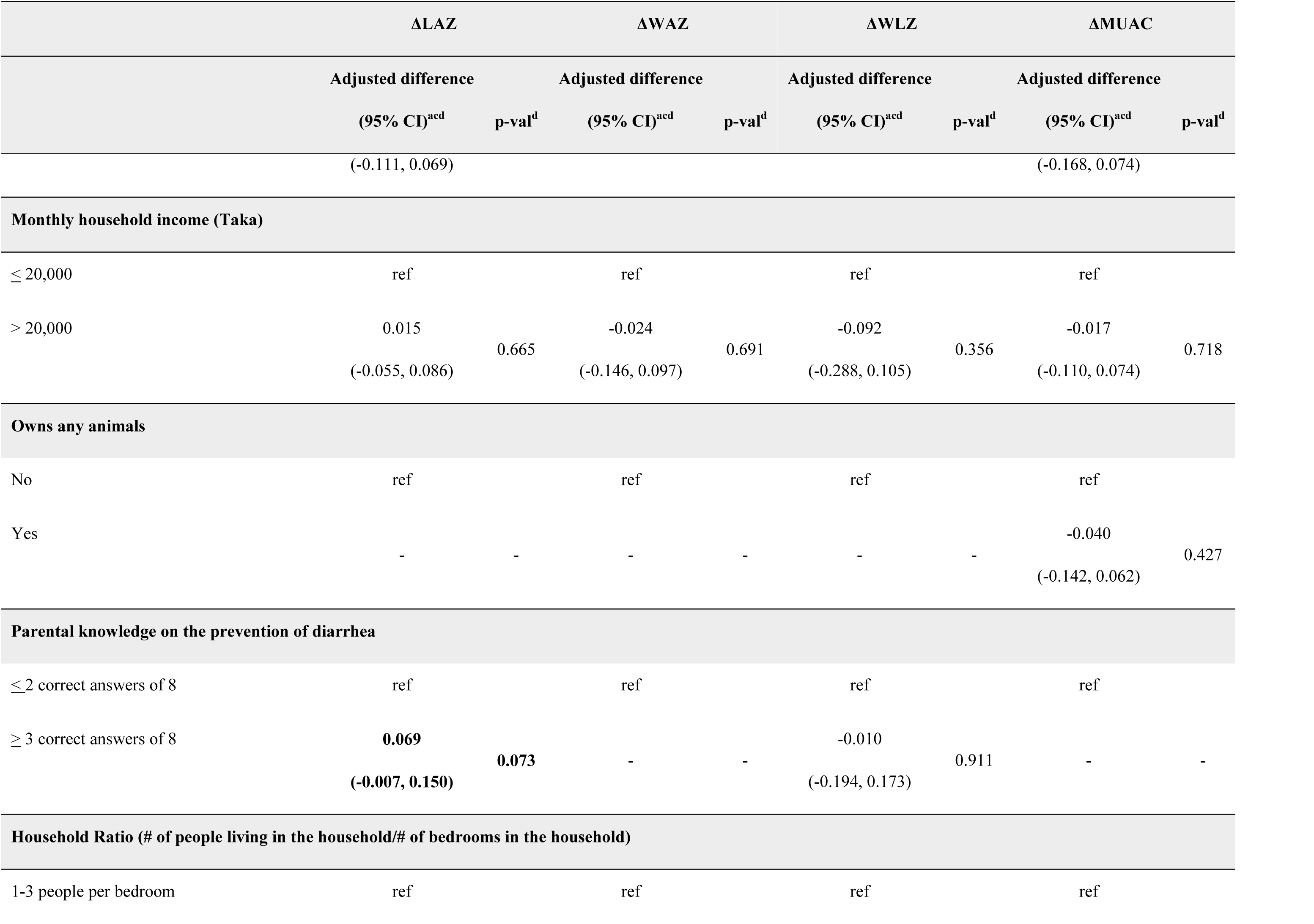
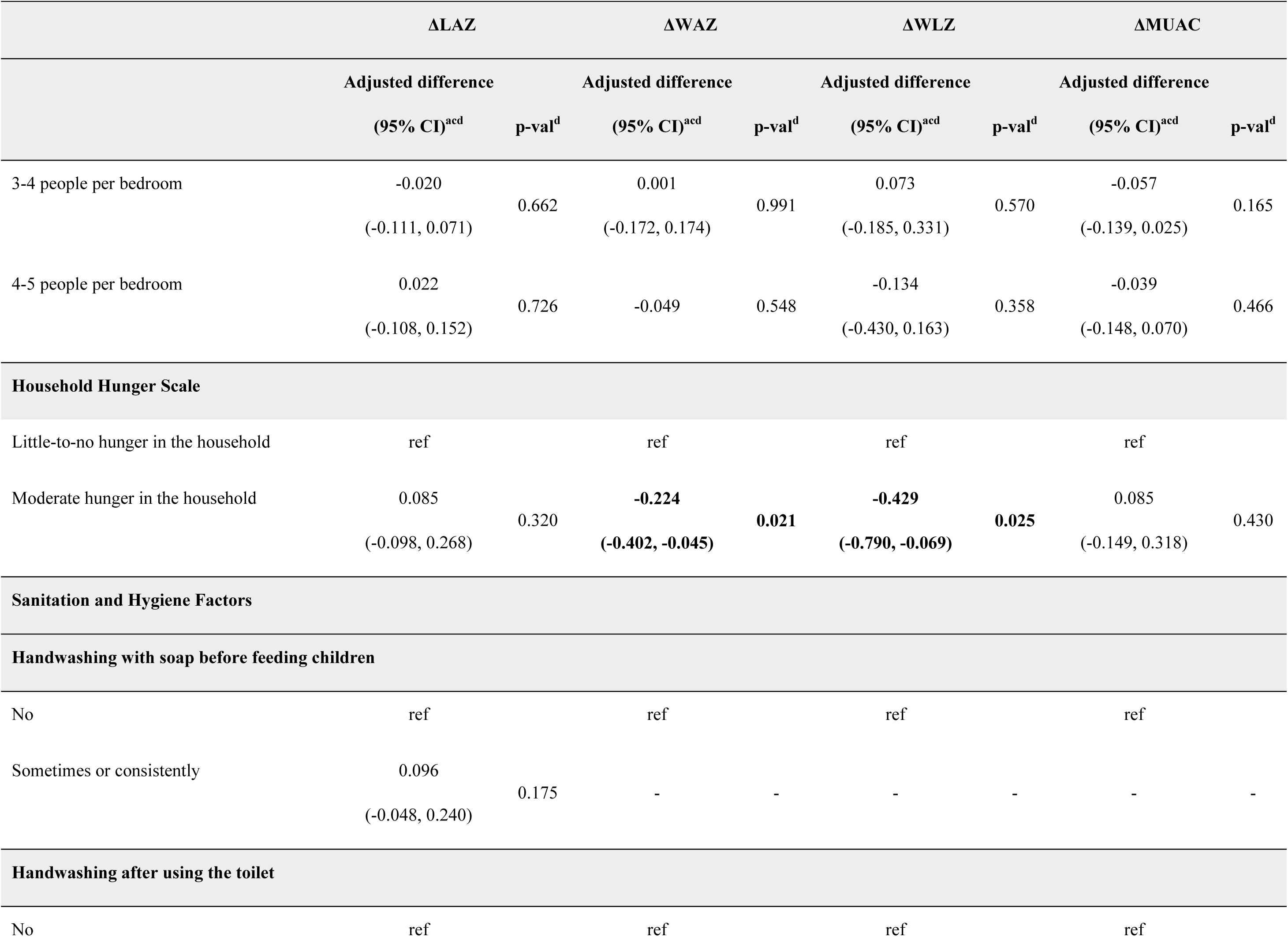
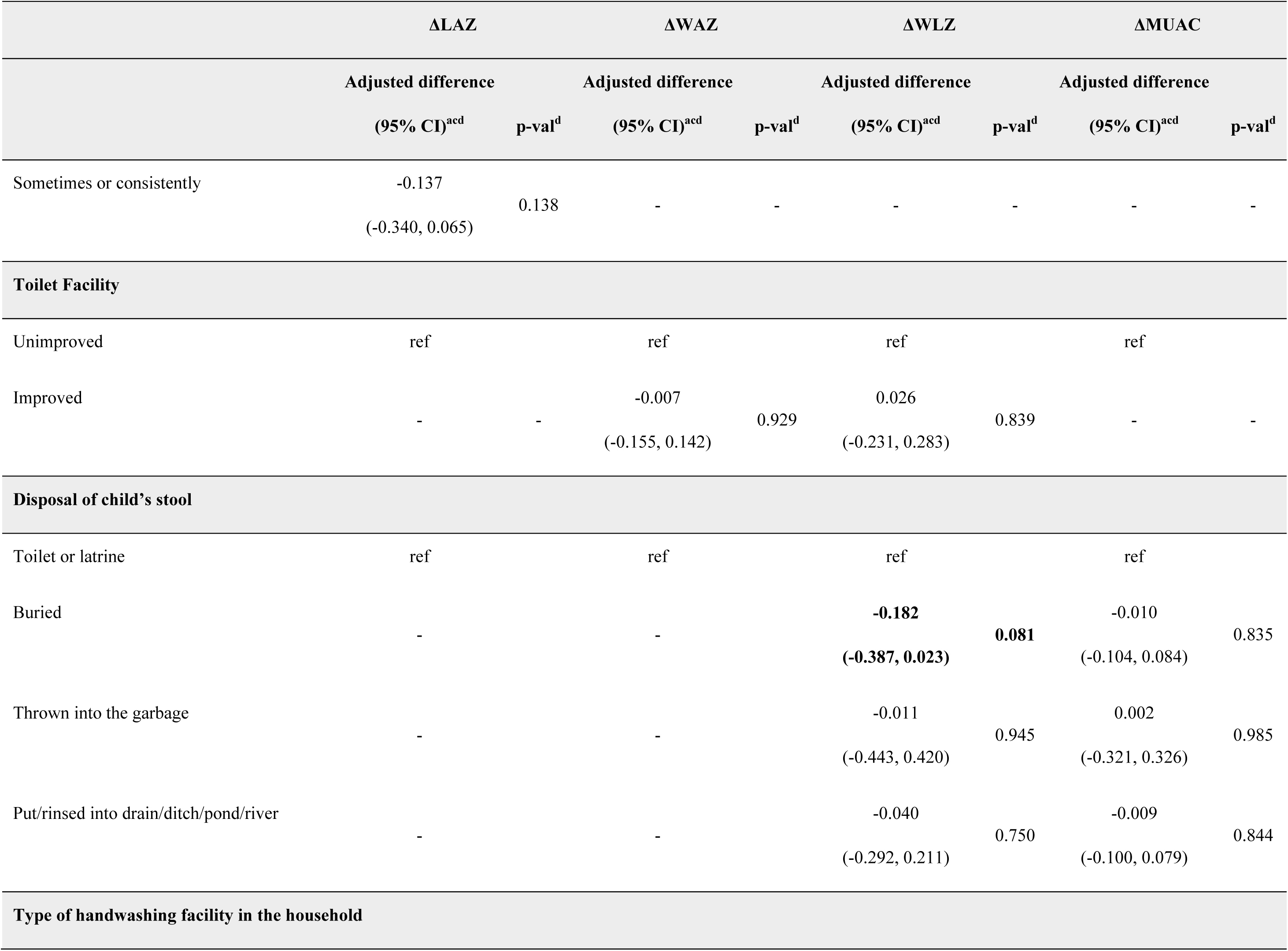
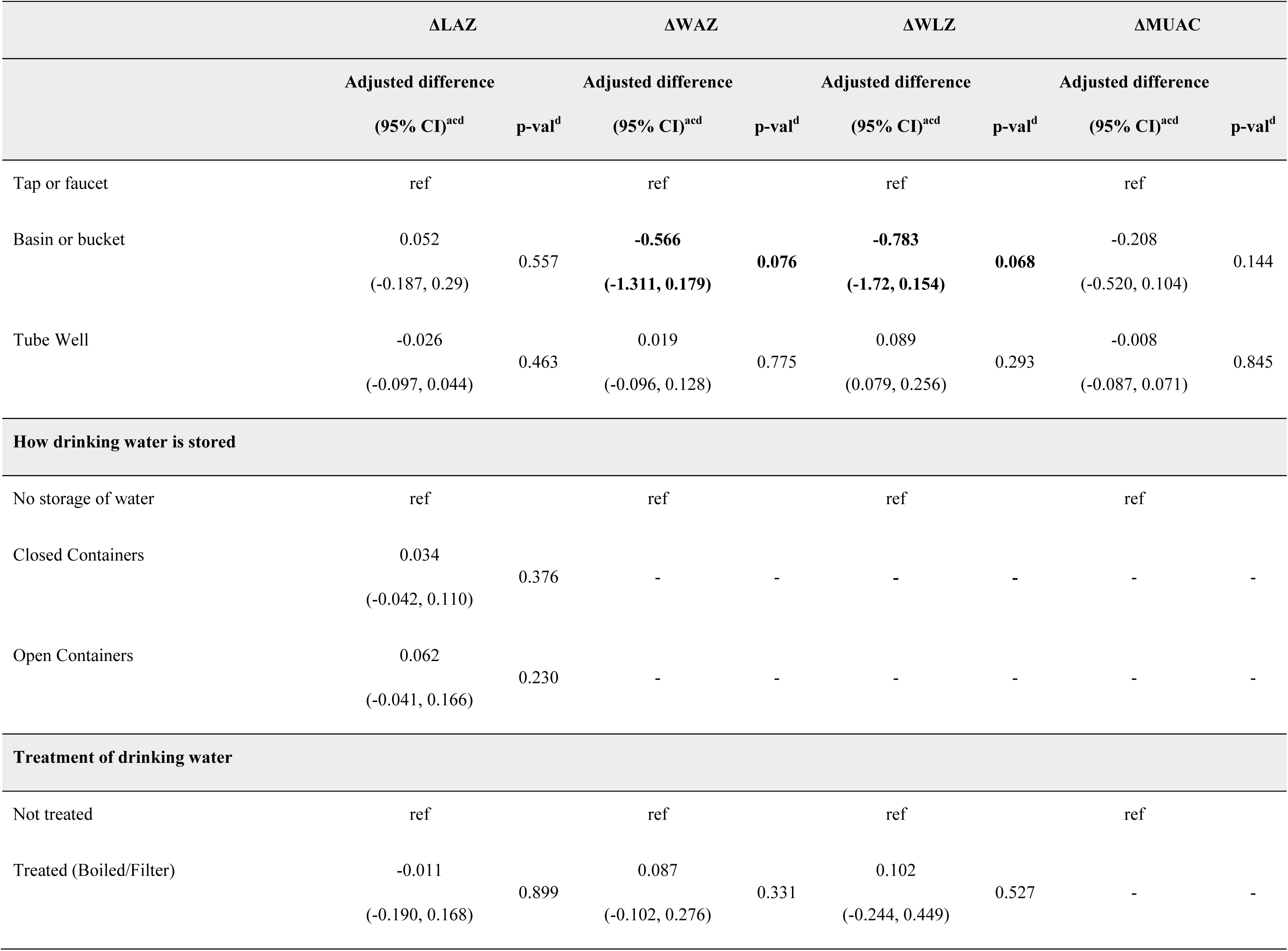
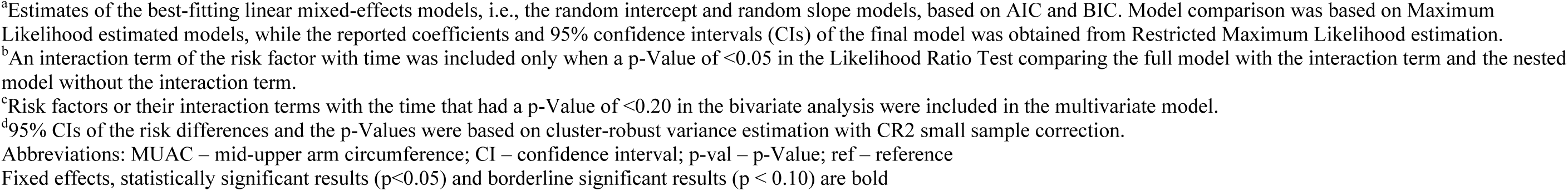
Risk factors associated with change in LAZ, WAZ, WLZ, MUAC during study follow-up among children under 24 months of age with watery diarrhea seeking care at Kumudini Hospital in Bangladesh.

#### 3.3.1 Clinical Factors

In bivariate models, clinical conditions such as diarrhea duration, vomiting, and hospitalization were associated with significant negative changes in anthropometric outcomes (Supplementary Table 1a). Children whose diarrheal disease required hospitalization at enrollment experienced a notable decrease in LAZ by 0.095 (p = 0.009, 95% CI: -0.164, -0.025). Children who had 7 days or more of diarrhea had a decrease in LAZ score of 0.085 (p = 0.090, 95% CI: -0.184, 0.014) at follow-up measurements than children who had diarrhea duration for zero to three days. Children who were hospitalized at enrollment for their diarrheal illness experienced lower MUAC scores at follow-up by 0.098 (p = 0.030, 95% CI: -0.185, -0.010).

#### 3.3.2 Sociodemographic Factors

In the bivariate analyses, age emerged as a significant demographic factor impacting nutritional outcomes (Supplementary Table 1b). Children aged 7 to 12 months had LAZ scores that were 0.161 higher (p = 0.008, 95% CI: 0.043, 0.278) compared to those younger than 6 months, while children aged 13 to 24 months showed a 0.113 increase in LAZ (p = 0.025, 95% CI: 0.015, 0.211) compared to those younger than 6 months at enrollment. However, age also modified the effect of time on weight and length measures; older children demonstrated a greater decline in WAZ, with children aged 7-12 months experiencing a 0.068 decrease (p = 0.022, 95% CI: -0.125, -0.010) and those aged 13-24 months showing a 0.096 decline (p < 0.001, 95% CI: -0.151, -0.042) relative to younger children. Similar trends were observed in WLZ, where older children had a greater decline over time compared to children under 6 months.

In the bivariate model, children in households with a higher occupancy ratio (3-4 people per bedroom) exhibited lower LAZ scores by 0.129 (p = 0.025, 95% CI: -0.241, -0.017) compared to those in less crowded homes. Higher income and maternal education levels were generally associated with improved scores across WAZ, WLZ, although specific values were not statistically significant after adjustment.

In the multivariate, adjusted model, children between the ages of 7 and 12 months experienced lower WAZ and WLZ scores when compared to children 1 to 6 months old. Their WAZ scores declined by 0.129 (p = 0.057, 95% CI: -0.262, 0.004) and their WLZ scores decreased by 0.242 (p = 0.049, 95% CI: -0.483, -0.0008) by the 90-day follow-up. Parental knowledge on the prevention of diarrhea impacted LAZ scores positively. Parents who had more knowledge in diarrhea prevention methods had LAZ scores that were greater by 0.069 (p = 0.073, 95% CI: -0.007, 0.150).

#### 3.3.3 Sanitation and Hygiene Factors

Sanitation and hygiene practices played a significant role in influencing growth outcomes (Supplementary Table 1c). Handwashing frequency and the type of handwashing facility were associated with notable changes in anthropometric measures. For example, children whose guardians practiced handwashing before feeding them had LAZ scores 0.155 higher (p = 0.009, 95% CI: 0.045, 0.266) than those who did not. However, the use of a basin or bucket for handwashing was associated with a decline in scores, with children using these facilities having lower WAZ scores by 0.618 (p <0.001, 95% CI: -0.713, -0.522), lower LAZ scores by 0.078 (p = 0.017, 95% CI: -0.142, -0.014), reduced WLZ (decrease of 0.740, p <0.001, 95% CI: -0.889, -0.590), and MUAC (decrease of 0.300, p <0.001, 95% CI: --0.359, -0.241). After adjustment, in the multivariate model, the type of handwashing facility did not impact the LAZ and MUAC measurements significantly but had borderline significance in the decrease of WAZ (decrease of 0.566, p = 0.076, 95% CI: -1.311, 0.179) and WLZ (decrease of 0.783, p = 0.068, 95% CI: -1.72, 0.154) measurements.

Access to improved sanitation also contributed positively to outcomes. Children in households with improved toilet facilities exhibited a 0.063 increase in WAZ (p = 0.017, 95% CI: 0.012, 0.115) and a 0.109 increase in WLZ (p = 0.007, 95% CI: 0.030, 0.188) scores over time.

#### 3.3.4 Food Security Factors

Food security and access to balanced meals were critical determinants of nutritional outcomes (Supplementary Table 1d). Limited access to protein-rich foods was associated with lower growth scores. Children from households that struggled to afford balanced meals containing fish or meat over the previous six months had significantly lower LAZ scores at follow-up by 0.132 (p = 0.042, 95% CI: -0.259, -0.005) compared to children with more consistent access to these foods.

The Household Hunger Scale provided further insights, indicating no cases of severe hunger and only a few instances of moderate hunger within the study population. Children from households with moderate hunger demonstrated declines in both WAZ and WLZ. Specifically, moderate hunger was associated with a 0.224 decrease in WAZ (p = 0.021, 95% CI: -0.402, -0.045) and a 0.429 reduction in WLZ (p = 0.025, 95% CI: -0.790, 0.069) at follow-up compared to children from households with little to no hunger. These findings underscore the role of food security in maintaining and improving nutritional outcomes among young children.

## 4. Discussion

This study provides valuable insights into the determinants of post-diarrhea growth outcomes among children under two in rural Bangladesh. Our findings demonstrate that growth outcomes following diarrheal episodes are influenced by a complex interplay of clinical, sociodemographic, sanitation, hygiene, and food security factors.

Children who were already malnourished at enrollment represent a particularly vulnerable subgroup that needs to be identified as quickly as possible as these children are at higher risk of persistent growth faltering and require targeted follow-up care to break the vicious cycle of undernutrition and recurrent infections. Our findings indicate that baseline malnutrition strongly predicts persistent or recurrent malnutrition. The vulnerability of this subgroup warrants the need of integrated interventions-combining nutritional support, healthcare access, and caregiver education- to address their specific needs.

The clinical burden of prolonged diarrhea emerged as a significant factor in growth declines, aligning with previous studies showing the detrimental impact of extended diarrheal episodes on nutritional recovery[20–22,25]. Children with longer diarrhea durations exhibited lower growth indicators, suggesting that the duration and severity of symptoms like vomiting, dehydration, fever, and anorexia might exacerbate malnutrition. This indicates the importance of sustained clinical support for children recovering from extended diarrheal periods to help mitigate the compounding effects of prolonged symptoms on growth outcomes. Effective post-discharge care may include support and follow-up for symptom management and nutritional supplementation, which could enhance recovery and resilience[26,27].

Younger children demonstrated more favorable growth responses in comparison to older children at enrollment, showing that age played an essential role in post-diarrhea growth outcomes. This may reflect the cumulative health burden carried by older children who have likely experienced repeated infections, weakening their resilience over time[28,29]. Additionally, household environment and sociodemographic factors, including family size, maternal education, and household income, were associated with variations in growth measures. These findings underscore that economic resources and educational background influence caregivers’ ability to maintain conditions that promote child growth. Socioeconomic interventions that enhance maternal knowledge of child health and nutrition could therefore help alleviate the cycle of infection and malnutrition in early childhood[30,31].

Sanitation and hygiene factors were also significantly associated with growth outcomes. Consistent handwashing practices among caregivers correlated with better growth outcomes, while shared or inadequate sanitation facilities were linked to an increased risk of growth faltering. The use of basins or buckets for handwashing was associated with poorer growth outcomes, suggesting that the type of handwashing facility may play a role in hygiene effectiveness[46,47]. Hygiene interventions were previously shown to significantly reduce diarrhea prevalence and improve child growth outcomes, further underscoring the importance of proper handwashing practices[35]. These findings reinforce the role of environmental sanitation in child health, aligning with literature that associates poor sanitation and hygiene with increased pathogen exposure and environmental enteric dysfunction[32,33,50]. Initiatives to improve community-level sanitation infrastructure, alongside education to reinforce proper hand hygiene, could substantially reduce the transmission of diarrheal pathogens and their impact on child growth[34,35]. However, the effectiveness of WASH (Water, Sanitation, and Hygiene) interventions has shown mixed results in some studies, with varying impacts on child growth and diarrhea prevalence[56]. This underscores the need for context-specific approaches that address local barriers and challenges to WASH implementation and adherence.

Furthermore, food security emerged as a critical factor, with households facing moderate hunger or limited access to protein-rich foods demonstrating poorer growth outcomes. Consistent with existing research, these findings highlight the importance of adequate dietary intake for growth recovery following infection and echoes the WASH Benefits study conclusion that nutrition interventions had stronger growth effects than WASH alone[36,37, 3]. Protein and other essential nutrients play an important role in immune function and growth, emphasizing the need for nutritional programs that prioritize food security. Programs that provide regular monitoring and supplementation, such as ready-to-use therapeutic foods (RUTF), have shown promise in improving growth outcomes in similar settings[56]. Integrating the Household Hunger Scale into routine assessments could help identify at-risk families and provide targeted support, such as RUTF, thereby enhancing child resilience against future growth challenges[38,39].

Some findings in this study were interesting and warrant further discussion. For instance, the continuous decline in length-for-age z-scores (LAZ) over the follow-up period contrasts with previous studies that have reported catch-up growth following diarrheal episodes[40,41]. A recent study by Brander et al. also found that linear growth faltering was strongly associated with diarrheal episodes, particularly in children with moderate-to-severe diarrhea, supporting our findings[20]. Additionally, the negative association between higher maternal education and LAZ scores was unexpected, as maternal education is typically associated with improved child health outcomes[42,43]. This finding may reflect confounding factors, such as working mothers having less time for childcare or differences in feeding practices and warrants further investigation.

This study has several strengths. The longitudinal design allowed for the examination of growth trajectories over time, providing insights into the dynamic nature of child growth following diarrheal episodes. The use of standardized anthropometric measurements and validated tools, such as the Modified Vesikari Severity Score and the Household Hunger Scale, enhances the reliability and comparability of the findings[25,38,53]. Additionally, the inclusion of a wide range of clinical, sociodemographic, and environmental factors provides a comprehensive understanding of the determinants of post-diarrhea growth outcomes. This study also contributes to the growing body of literature on the interplay between diarrhea, nutrition, and child growth in low-resource settings, offering valuable evidence for the design of targeted interventions[48,49].

This study has limitations. Our study only includes children seeking care from the hospital and thus miss the opportunity to analyze community diarrhea cases. Our study setting represents a rural community of Bangladesh that may not represent other populations in the country. Although very few, our exclusion criteria did not allow for severely malnourished (WLZ < -3) children presenting to the study hospital to join the study. The low number of children experiencing a transition in their undernutrition status did not allow us to study the independent association of the risk factors of interest with these binary outcomes.

## 5 Conclusion

In conclusion, this study highlights the multifactorial nature of child growth and the critical role of age, household practices, environmental factors, and clinical symptoms in influencing nutritional outcomes after diarrhea incidence. Interventions that focus on improving hygiene, sanitation, and access to healthcare, alongside age-specific nutritional education, are essential for supporting child growth and development. By addressing these determinants, public health initiatives can effectively combat malnutrition and enhance the overall well-being of children.

## Data Availability

All data produced in the present work are contained in the manuscript

## 6 Conflict of Interest

The authors declare no conflict of interest.

## 7 Author Contributions

Conceptualization, S.C. and A.S.G.F.; Methodology, S.C., A.S.G.F., S.D., J.P., F.T., M.R., M.A., D.A.S., and B.R., and A.A.H.; Investigation, A.S.G.F., S.C., S.D., F.T., M.A., M.R., B.R., J.L., F.N., F.A., and E.S.; Writing—original draft preparation, J.L., Y.Z., J.P., Writing—review and editing, S.C., A.S.G.F., S.D., F.T., M.R., T.A., M.A., F.N., E.S., B.R., F.A., A.A.H., Y.Z, D.A.S., and J.L.; Funding acquisition, S.C. All authors have read and agreed to the published version of the manuscript.

## 8 Funding

This study was funded by the National Institute of Allergy and Infectious Diseases of National Institute of Health, R01AI153399 (SC). The content of this article is solely the responsibility of the authors and does not necessarily represent the official views of the NIH, NIAID.

## 9 Acknowledgments

The authors thank the participants, and their caregivers enrolled in the INSIGHT study. We thank Sampa Bakali, Afrin Sultana Ratna, Taposhi Das, Shikha Sultana, Ayesha Akter, Suchi Roy, Lovely Akter, Shamim Al Mamun, Razib Sikdar, Shaniram Das and all the other staff of the INSIGHT study for their hard work and invaluable contributions. We acknowledge the contribution of icddr,b’s core donors, including the Government of the People’s Republic of Bangladesh, Global Affairs Canada (GAC), Canada, for their continuous support and commitment to icddr,b’s research efforts.

## Notes

### Competing Interest Statement

The authors have declared no competing interest.

### Clinical Protocols

https://doi.org/10.3390/microorganisms12081677

